# Fetoscopic Endoluminal Tracheal Occlusion with Smart-TO balloon: efficacy of removal and safety

**DOI:** 10.1101/2022.08.17.22278918

**Authors:** Nicolas Sananès, David Basurto, Anne-Gaël Cordier, Caroline Elie, Francesca Russo, Alexandra Benachi, Jan Deprest

**Affiliations:** Department of Maternal Fetal Medicine, Strasbourg University Hospital, France; INSERM 1121 ‘Biomaterials and Bioengineering’, Strasbourg University, France; MyFetUZ Fetal Research Center, Department of Development and Regeneration, Cluster Woman and Child, Biomedical Sciences, KU Leuven, Belgium; Department of Maternal fetal Medicine, Antoine–Béclère Hospital - Paris–Saclay University, Clamart, France; Clinical Research Unit / Clinical Investigation Center, Necker-Enfants malades Hospital, Paris, France; Clinical Department of Obstetrics and Gynaecology, University Hospitals Leuven, Belgium; Institute for Women’s Health, University College London, UK

## Abstract

**Introduction:** One of the drawbacks of fetoscopic endoluminal tracheal occlusion (FETO) for congenital diaphragmatic hernia is the need for a second invasive intervention to reestablish airway patency. The “Smart-TO” (Strasbourg University-BSMTI, France) is a new balloon for FETO, which spontaneously deflates when positioned near a strong magnetic field, e.g., generated by a magnetic resonance image (MRI) scanner. Translational experiments have demonstrated its efficacy and safety. We will now use the Smart-TO balloon for the first time in humans. Our main objective is the efficacy of prenatal deflation of the balloon by the magnetic field generated by an MRI scanner.

**Material and methods:** This is a phase-I study conducted in the fetal medicine units of Antoine–Béclère Hospital, France, and UZ Leuven, Belgium. Conceived in parallel, protocols were amended by the local Ethics Committees, resulting in some minor differences. This trial is a single-arm interventional feasibility study. Twenty (France) and 25 (Belgium) participants will have FETO with the Smart-TO balloon. Balloon deflation will be scheduled at 34 weeks or earlier if clinically required. The primary endpoint is the successful deflation of the Smart-TO balloon after exposure to the magnetic field of an MRI, assessed through ultrasound immediately after MRI-exposure. The secondary objective is to report on the safety of the balloon. The percentage of fetuses in whom the balloon is deflated after exposure will be calculated with its 95% confidence interval. Safety will be evaluated by reporting the nature, number, and percentage of serious unexpected or adverse reactions.

**Conclusion:** This phase-I study may provide the first evidence of the potential to reverse the occlusion by Smart-TO and free the airways non-invasively, as well a safety data.

## INTRODUCTION

### Background

Congenital diaphragmatic hernia (CDH) is a birth defect characterized by failed closure of the diaphragm. This enables abdominal viscera to herniate into the thoracic cavity, leading to hypoplastic lungs and impaired lung vasculature [1]. Fetoscopic Endoluminal Tracheal Occlusion (FETO) increases fetal lung volume and therefore can improve survival in selected fetuses with CDH. Recently two parallel randomized controlled trials in fetuses with isolated left-sided CDH with severe and moderate pulmonary hypoplasia respectively were concluded [2, 3]. In severe hypoplasia the balloon was inserted early (27^+0^ to 29^+6^ weeks’ gestation) and FETO improved survival from 15% to 40% (Table 1) [3]. A comparable improvement in survival (20% to 42%) was achieved in fetuses with severe right-sided CDH [4]. In moderate hypoplasia, the balloon was inserted later (30^+0^ to 31^+6^ weeks’ gestation) in an effort to reduce the risks of very preterm birth. In that study, FETO improved survival from 50% to 63%, but the difference in survival was not statistically significant [2]. Analysis of the pooled data from the two randomized trials demonstrated that FETO increases survival in both severe and moderate disease (Table 1), but the observed lesser effect in the moderate group is most likely a mere consequence of the delayed insertion of the balloon in moderate hypoplasia[5].

**Table 1:** Outcomes of fetuses diagnosed with isolated congenital diaphragmatic hernia (CDH) in the prenatal period, either left or right-sided, expectantly managed during pregnancy or having tracheal occlusion within the “Tracheal Occlusion to Accelerate Lung Growth” (TOTAL) trial or and a large study on right-sided CDH under the same management protocol.

| Side, severity | Criteria severity on ultrasound | Survival to discharge |  | RR (95% CI) |
| --- | --- | --- | --- | --- |
|  |  | Expectant | FETO |  |
| Isolated left sided CDH – TOTAL trials |  |  |  |  |
| TOTAL<br>severe[3] | O/E LHR <25.0%<br>Irrespective of liver position | 6/40<br>(15%) | 16/40<br>(40%) | 2.67<br>(1.22-6.11) |
| TOTAL<br>moderate[2] | O/E LHR 25.0-34.9%, any liver position<br>O/E LHR 35.0-44.9% & liver into chest | 49/98<br>(50%) | 62/98<br>(63%) | 1.27<br>(0.99-1.63) |
| Isolated left sided CDH – Pooled analysis TOTAL data |  |  |  |  |
| Late<br>insertion | O/E LHR 0.0-34.9%, any liver position<br>O/E LHR 35.0-44.9% & liver into chest | 55/142<br>(39%) | 79/145<br>(54%) | A OR: 1.78<br>(1.05-3.01) |
| Early<br>insertion | O/E LHR 0.0-34.9%, any liver position<br>O/E LHR 35.0-44.9% & liver into chest |  |  | A OR: 2.73<br>(1.15-6.49) |
| Isolated right sided CDH |  |  |  |  |
| Severe[4] | O/E LHR <50%<br>Irrespective of liver position | 7/34<br>(20%) | 53/125<br>(42%) | 2.84<br>(1.15-7.01) |

Abbreviations: RR, risk ratio; CI, confidence interval; O/E LHR, observed-to-expected lung-to-head ratio; A, adjusted; OR, odds ratio.

An adverse side-effect of FETO is that it increases the risk for iatrogenic preterm membrane rupture and preterm birth [6, 7]. In the TOTAL trials, that risk was inversely related to the gestational age at the insertion of the balloon [5]. Although the trials did not demonstrate any obvious differences between the FETO and control groups in prematurity-related complications, they were not powered to study differences in these secondary outcomes. Long-term outcomes will have to further elucidate that, but it would seem logical to expect a measurable effect of prematurity when large numbers are available.

A second disadvantage of the current procedure is the need for a second intervention to reverse the occlusion and re-establish airway patency. Balloon removal is scheduled electively at 34 weeks, or earlier if required. Reversal of the occlusion is preferentially performed at least 24 hours before birth, as that seems associated with an increased survival [2, 8-10]. Reversal is at present an invasive procedure that can be performed by either ultrasound-guided puncture, fetoscopy, or, less ideal, while the fetus is maintained on placental circulation or at birth after vaginal delivery [10]. Airway re-establishment requires a specialist team familiar with the procedure and that is available 24/7 [10]. In a large series, 28% of balloon removals were in an emergency setting [10]. The only neonatal deaths that occurred, were when balloon reversal was attempted in centers without experience or that were unprepared [10]. Even in experienced centers balloon removal can fail, as observed in the TOTAL trial [2]. Also, patients may be non-compliant and move away from the fetal surgery center [2]. The second procedure inherently adds risks for the mother and fetus. These can be directly procedure-related, but also indirectly, by increasing the risk for membrane rupture later on [10]. In conclusion, the occlusion period is a serious burden on *patients* who are requested to stay close to the FETO center until balloon removal, as well as for the *fetal surgery centers* because of the need for permanently available staff. All these conditions, limit the acceptability of FETO as being practiced today.

The University of Strasbourg, France, in partnership with BS Medical Tech Industry (BS-MTI), Niederroedern, France, developed an alternative occlusion device, referred to as “Smart-TO” [11]. Compared to the currently used Goldbal2® (Balt, Montmorency, France) balloon, the Smart-TO balloon has identical dimensions in its inflated state and is made of the same material (latex). Around the balloon neck, there is a metallic cylinder and inside a magnetic ball, which together act as a valve. Deflation occurs under the influence of a strong magnetic field, which is present around any clinical MRI machine. For that, it is sufficient for the pregnant woman to walk around the MRI machine. This enables non-invasive, externally controlled balloon deflation. The Smart-TO balloon been tested preclinically by BS-MTI (the manufacturer), University of Strasbourg, Simian Laboratory Europe and the KU Leuven. In-vitro tests including permeability, occlusion, and deflation in a simulated environment were performed by BS-MTI (unpublished data). Deflation tests were performed using a mannequin in a simulated “in-utero” environment, with the fetus and the mother in different positions and heights. In that experiment, deflation was successfully achieved regardless of the fetal position and the exact level of the fetus from the ground [12]. In vivo animal tests included the demonstration of similar lung growth and short-term tracheal side effects as the Goldbal 2 balloon in fetal lambs [12] [13]. In the latter experiment, fetal lambs expelled the Smart-TO balloon following exposure to the fringe field of a 3T MRI. Finally, feasibility of balloon insertion, persisting occlusion until reversal, and spontaneous expulsion of the Smart-TO balloon was confirmed in non-human primates [11]. Therefore, this novel medical device should now be evaluated in a phase I or first-in-human study. For that purpose we designed two parallel studies, one at Antoine–Béclère Hospital Paris–Saclay University, Clamart, France referred to as “Smart-FETO”, and one at the University Hospitals Leuven (UZ Leuven), Belgium, referred to as “Smart-Removal”. Conceived in parallel, protocols were amended by the local Ethics Committee on Clinical Studies or its equivalent, resulting in a limited number of differences (Table 2).

**Table 2:** Inclusion criteria and outcome measurements in both studies. Differences are displayed in bold.

| Smart-FETO (Paris) | Smart-Removal (Leuven) |
| --- | --- |
| Inclusion criteria |  |
| - Patient can consent, has 18 years or more, and **has medical insurance**<br>- Singleton pregnancy with a fetus with an isolated left CDH with severe or moderate lung hypoplasia | - Patient can consent, has 18 years or more<br>- Singleton pregnancy with a fetus with an isolated left CDH with severe or moderate lung hypoplasia or **right CDH with severe lung hypoplasia** |
| (Co-)primary endpoint |  |
| - Balloon deflation rate after MRI exposure (by ultrasound).<br>- **Balloon expulsion from the fetal airways (by postnatal chest X-ray)** | - Balloon deflation rate after MRI exposure (by ultrasound). |
| Secondary endpoints |  |
| <u>Prenatal</u><br>- Spontaneous balloon deflation prior to MRI exposure (by ultrasound)<br>- Lung growth (O/E LHR).<br>- **Balloon height and width** (ultrasound).<br>- Gestational age at membrane rupture<br><u>Postnatal:</u><br>- Gestational age at delivery<br>- **Localisation of the balloon by postnatal chest X-ray of the newborn**<br><u>Neonatal:</u><br>- Survival at discharge from the NICU<br>- Survival at 6 months<br>- Need for oxygen supplementation at 6 months<br><u>Adverse events:</u><br>- Any adverse event, either in the mother or the fetus or newborn, at whatever time point between insertion and discharge from the neonatal unit | <u>Prenatal</u><br>- Spontaneous balloon deflation prior to MRI exposure (by ultrasound).<br>- Lung growth **2 weeks after FETO** (O/E LHR).<br>- Gestational age at membrane rupture<br><u>Postnatal:</u><br>- Gestational age at delivery<br>- Balloon expulsion from the fetal airways (by postnatal chest X-ray)<br>- **Localisation of the balloon either by (1) direct visualization within the amniotic fluid, membranes or placenta, (2) postnatal chest X-ray of the newborn, and (3) ultrasound of the postpartum uterus.**<br><u>Neonatal:</u><br>- Survival at discharge from the NICU<br>- **Tracheal diameter on first postnatal chest X-ray**<br>- **Assessment for any local side effects of the balloon (signs or symptoms of tracheomegaly and /or tracheomalacia)**<br><u>Adverse events:</u><br>- Any adverse event, either in the mother or the fetus or newborn, at whatever time point between insertion and discharge from the neonatal unit |
| Sample size |  |
| n=20 for a 95% CI with a lower boundary of 83%[14]. | n=25 for a 95% CI with a lower boundary of 85%[14] and an expected loss rate of 8% (n=2) due to the need for removal on placental circulation or spontaneous balloon deflation. |

Abbreviations: CDH, congenital diaphragmatic hernia; MRI, magnetic resonance image; O/E LHR, observed to expected lung to head ratio; CI, confidence interval.

### Objectives and hypotheses

The main objective of the study is to demonstrate the ability to consistently deflate the balloon prenatally by the magnetic fringe field generated by a clinical MRI scanner, and that it will be expelled from the airways. Secondary objective is to report on the safety of the balloon. We hypothesize that there will not be any serious adverse effects directly related to the Smart-TO balloon itself. Other objectives include assessment of prematurity, preterm premature rupture of membranes, lung growth, neonatal survival, and the need for oxygen supplementation at discharge from the hospital.

## DESIGN PLAN

### Study type

This clinical trial is a single-arm interventional feasibility study. Eligible consecutive consenting women will have FETO with the Smart-TO balloon.

### Setting

The trial is conducted at two centers i.e., the Antoine–Béclère Hospital - Paris–Saclay University, Clamart, France, and the University Hospitals Leuven, Belgium

## SAMPLING PLAN

### Existing data

Both trials have been registered prior to their inception (ClincalTrial.gov NCT04931212 and NCT05100693). The first inclusion in France was on August 4^th^,2021, and in Belgium on September 10^th^,2021.

### Recruitment

Recruitment of participants will be at the latest one day before planned balloon placement.

### Inclusion Criteria

- Patient aged 18 years or more and who can consent,
- Singleton pregnancy with a fetus with an isolated congenital diaphragmatic hernia (i.e., no additional major structural malformation nor genetic abnormality)
- Eligible for FETO, i.e. having severe pulmonary hypoplasia defined as, in left-sided cases, an observed-to-expected ‘lung-to-head ratio’ (O/E LHR) <25% irrespective of the liver position, or moderate pulmonary hypoplasia defined as O/E LHR 25-34.9% irrespective of the liver position or O/E LHR 35-44.9% with liver herniation, and, in UZ Leuven, fetuses with right-sided CDH with severe hypoplasia (O/E LHR < 50%).

### Exclusion Criteria

- Maternal contraindication to fetoscopy
- Preterm premature rupture of the membranes (PPROM) or any condition strongly predisposing to PPROM or premature delivery
- Patient does not consent to stay close to the FETO center during the occlusion period.

### Sample size

Independent sample size calculation has been performed in both centers. In Paris (France) we hypothesized that for a 100% deflation and expelling rate, the estimated number of patients is 20 patients in order to achieve a 95% confidence interval (CI) with a lower boundary of 83% (calculation of the CI of a proportion using the exact method) [14]. In Leuven (Belgium), the estimated number is 23 patients, in order to achieve a 95% CI with a lower boundary of 85% [14]. The theoretical possibility of spontaneous balloon deflation, or the impossibility to expose the patient to MRI at the time of balloon removal (e.g., in an emergency requiring removal on placental circulation) was considered as possible (n=2), so that a total of 25 patients are to be recruited.

## VARIABLES

### Measured variables

These include administrative data, data on the index pregnancy, characteristics of the fetus, on the FETO procedure, follow-up ultrasound measurements, balloon removal, delivery, and the neonatal follow-up period until discharge from the neonatal intensive care unit (NICU) (Table 3).

**Table 3:**
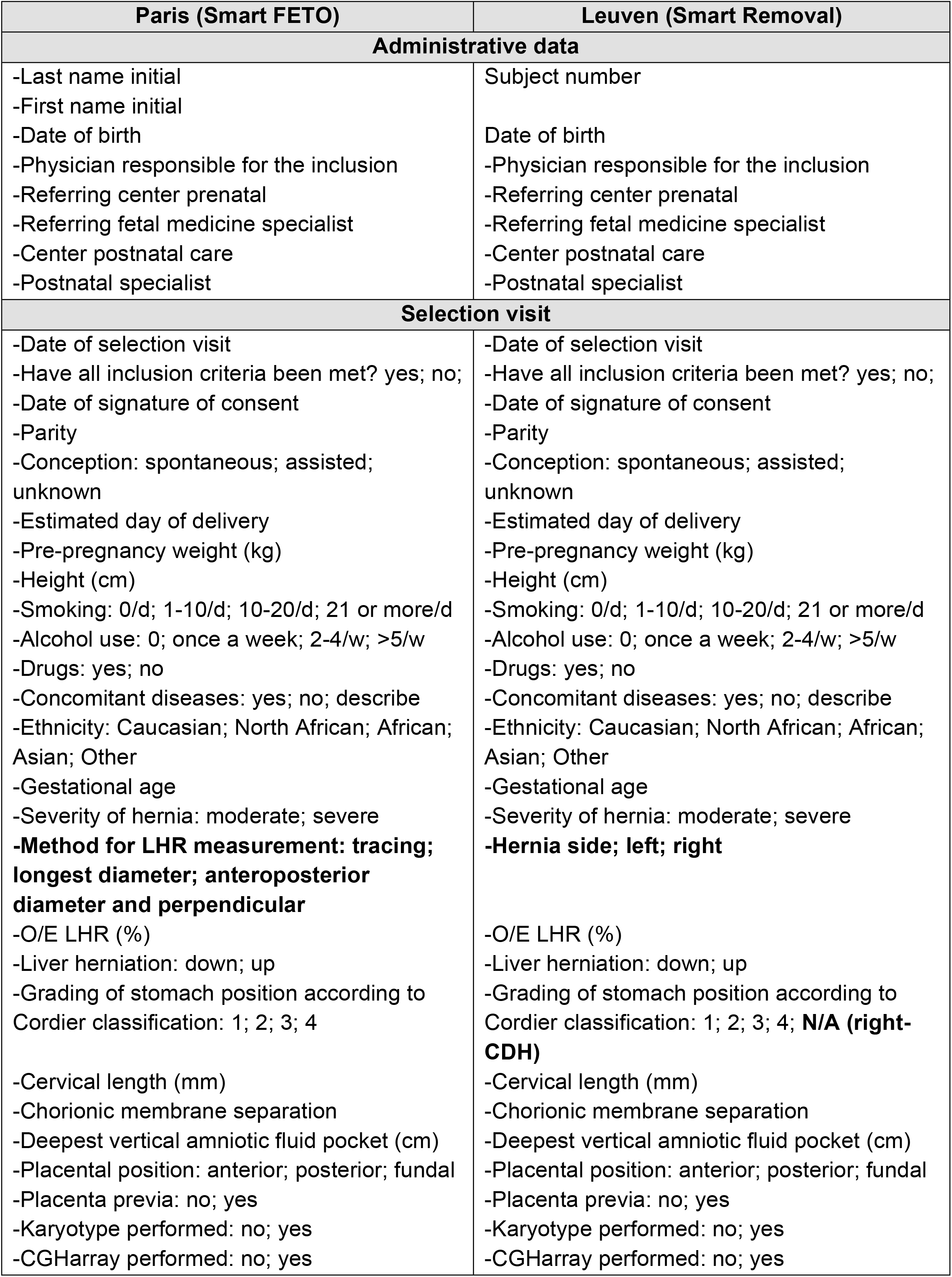
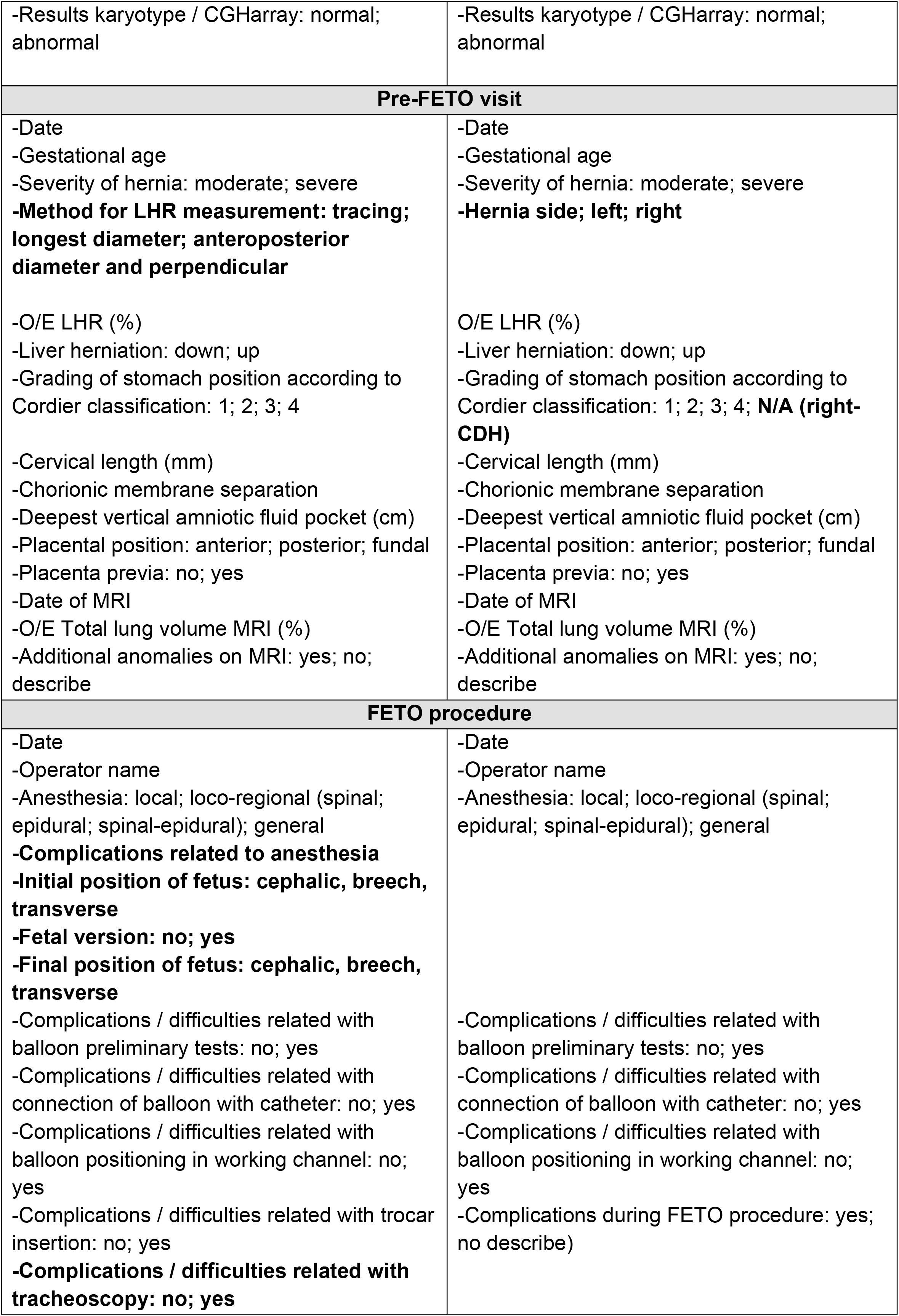
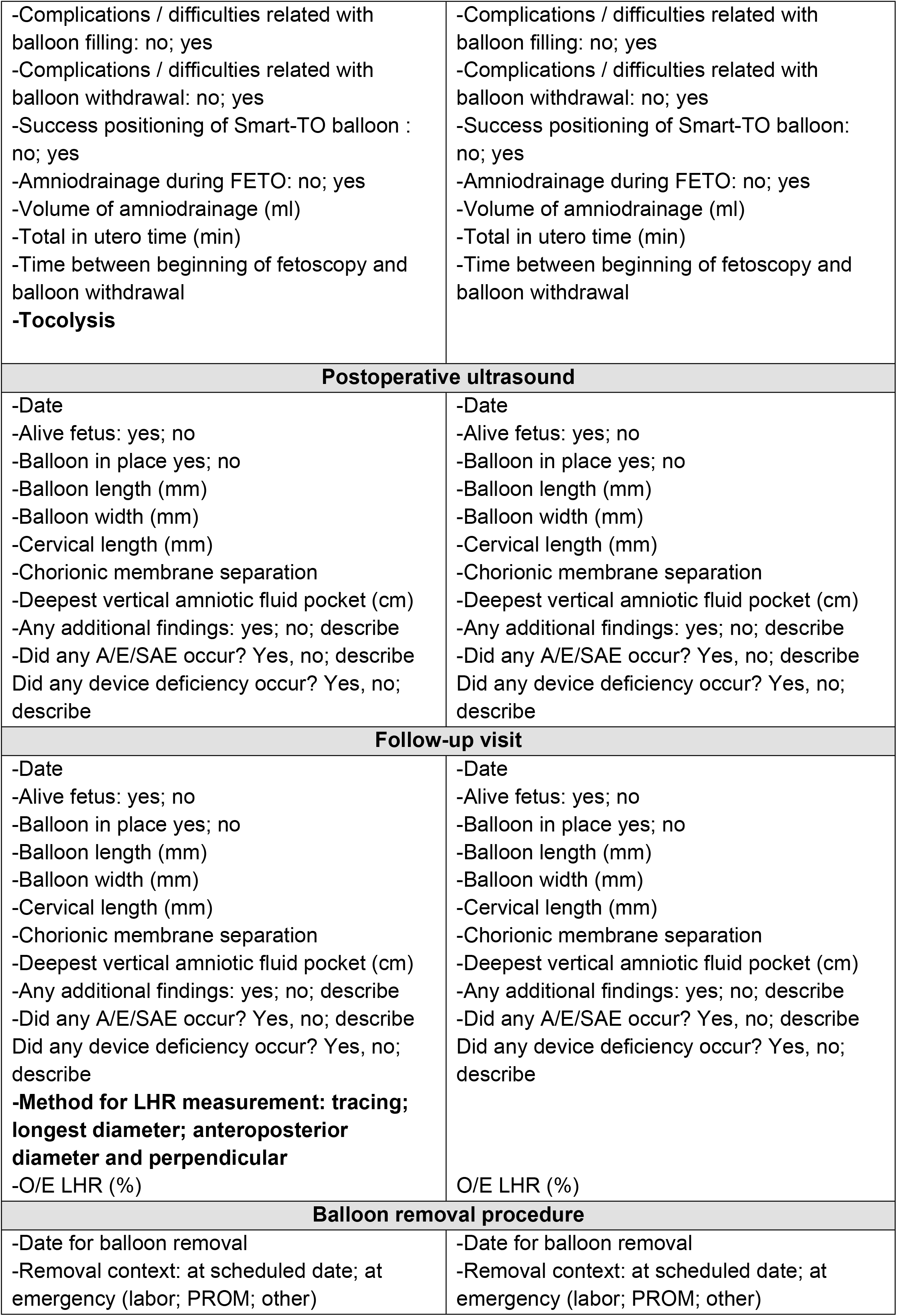
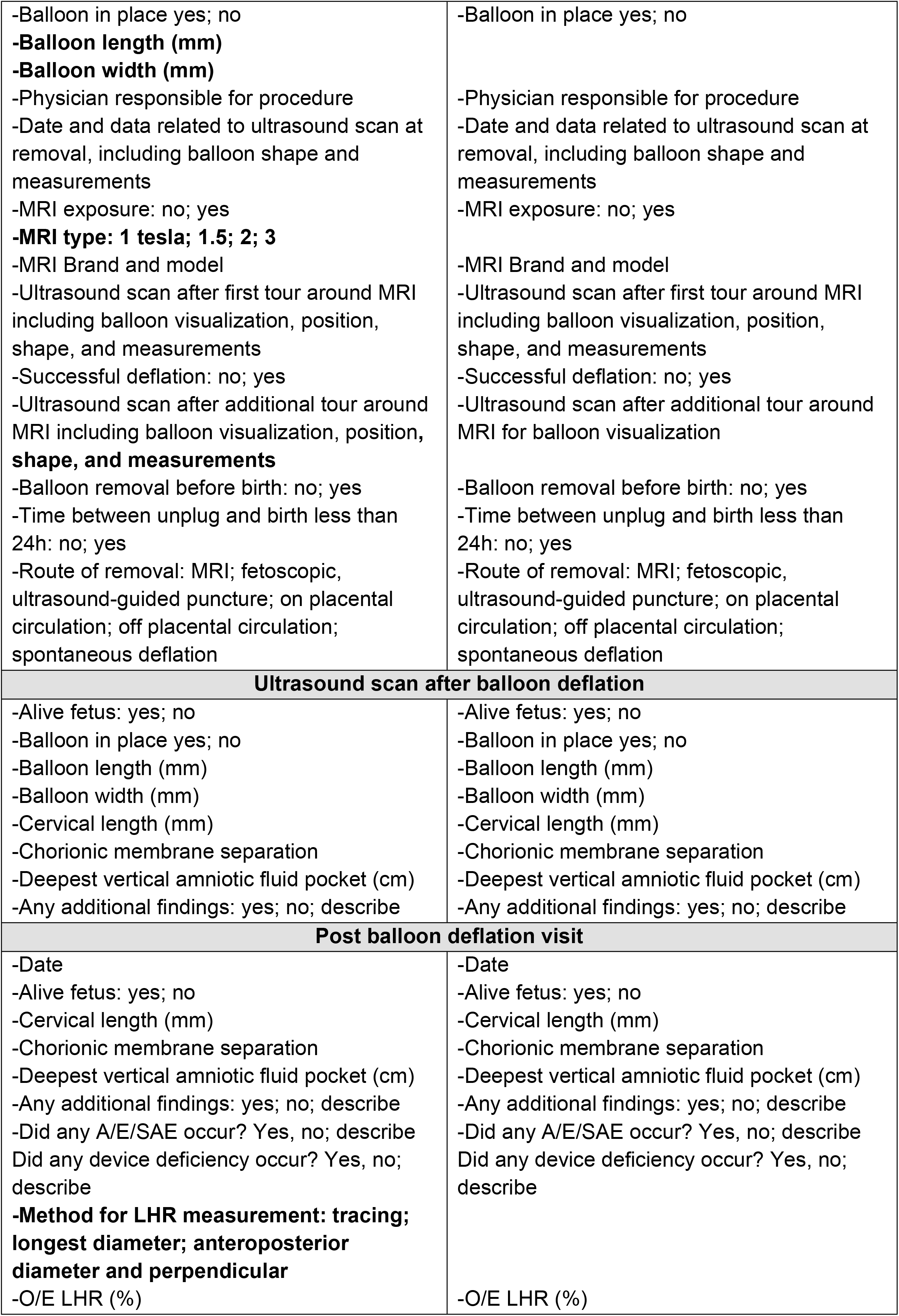
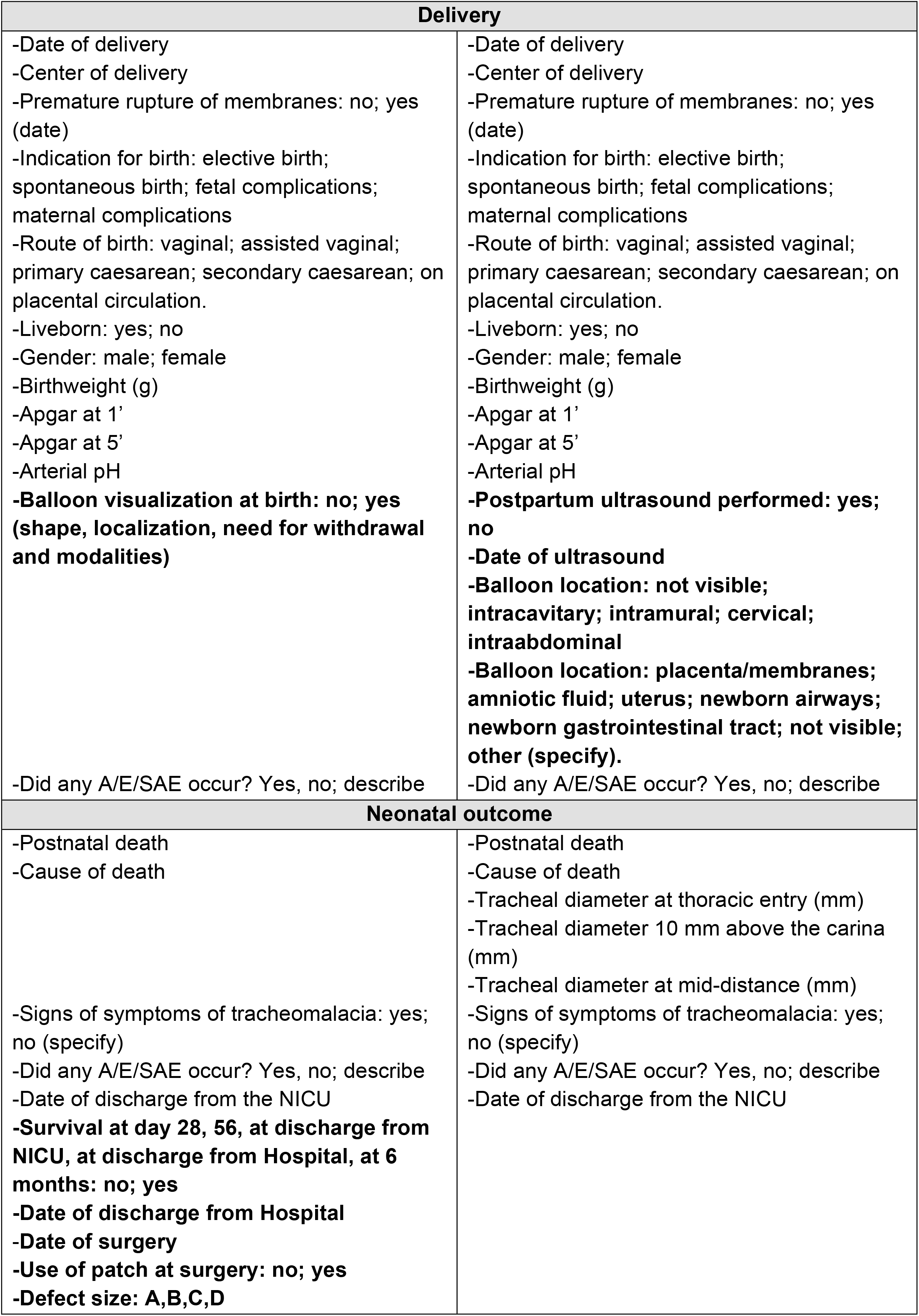
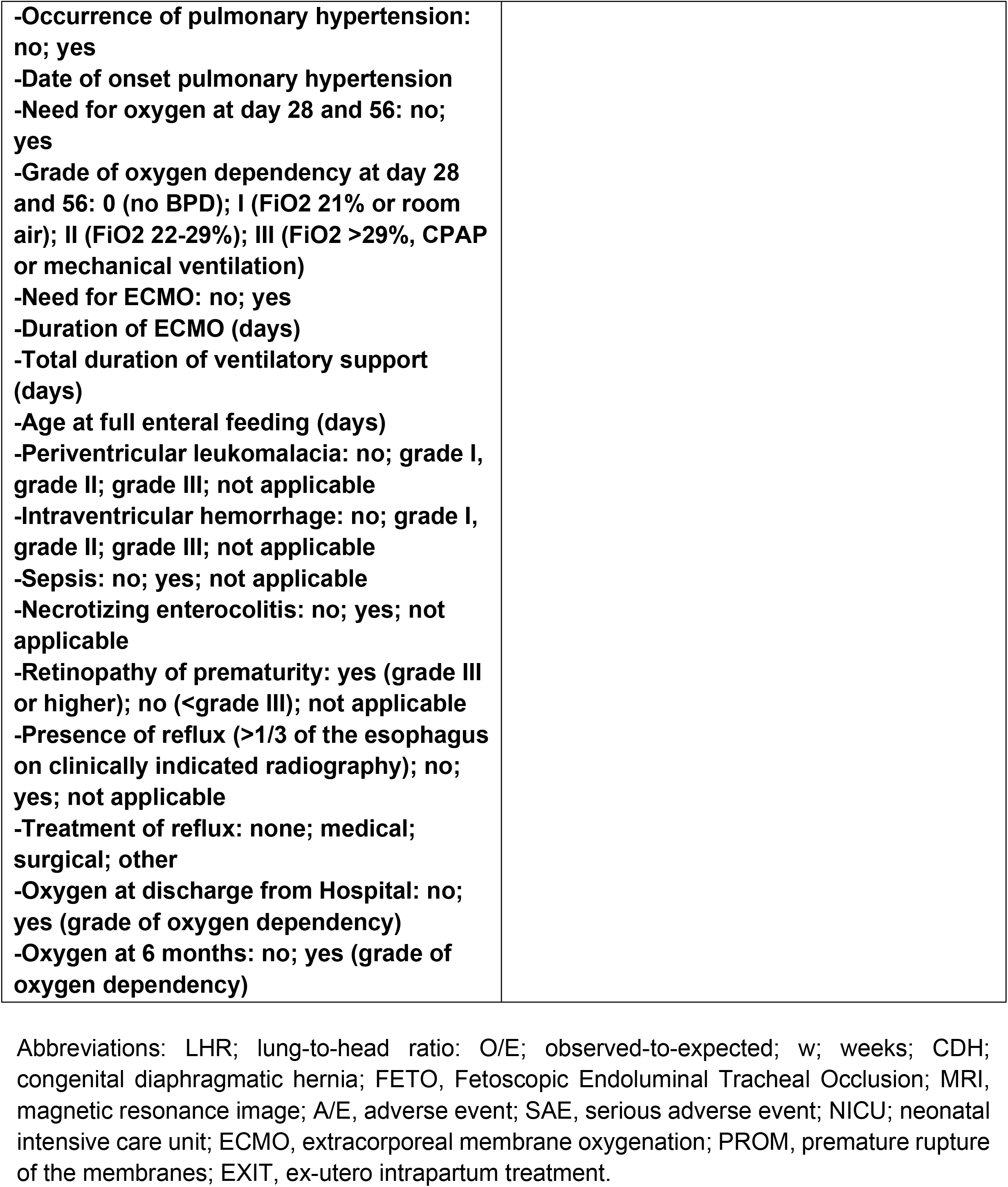
List of variables from both studies. Differences are displayed in bold.

### Primary endpoint

The primary endpoint is the successful deflation of the Smart-TO balloon after exposure to the fringe field of the MRI, assessed through ultrasound immediately after MRI exposure. For France, a co-primary endpoint is the expelling of the Smart-TO balloon from the airways, as documented by a X-ray of the neonatal chest at birth.

Secondary endpoints are displayed in table 2.

## STATISTICAL ANALYSIS PLAN

Quantitative data will be expressed as median and inter-quartile-range (IQR), qualitative data will be expressed as numbers and percentages. The percentage of fetuses in whom the balloon deflated at exposure and the percentage of fetuses that expelled the balloon from the fetal airways will be calculated with its 95% confidence interval using the binomial method [14]. Safety will be evaluated by reporting the nature, number, and percentage of serious unexpected or adverse reactions. We expect there will be no missing data for the primary outcome variable. There will be no imputation of missing data for secondary outcomes. A sensitivity analysis will be performed assuming the worst possible outcome for missing data.

## INTERVENTION

### FETO

The FETO procedure will be performed as earlier described [15]. Regarding the Smart TO use:

- The catheter system is introduced in the sheath of the endoscope and back loaded with the Smart-TO balloon. The balloon is then tested by inflation with 0.7 mL of sterile saline and deflated with its proper stylet, following which the latter is withdrawn.
- The balloon is positioned between the carina and the vocal cords, inflated with 0.7 mL sterile saline, and detached by the combination of gentle traction of the delivery system and counter pressure with the endoscope.

### Reestablishment of the fetal airways

The Smart-TO deflation protocol is displayed in Supplementary Video 1.

- The patient is positioned in front of the MRI, her abdomen facing the front of the tunnel of the machine.
- The patient walks (or is strolled) around the machine while staying as close as possible to the machine.
- When approaching the rear of the tunnel, the patient positions herself in the middle of it, facing the tunnel and makes a short stop.
- Then she continues to walk (or being strolled) around the MRI while staying as close as possible to the machine
- Once she has completed the turn, she can leave the MRI room.

Ultrasound is then performed independently by two experienced sonographers, to assess balloon deflation. When inflated, the balloon is easily visible on ultrasound as an anechoic structure. Balloon deflation will be indicated by visualization of the balloon on ultrasound before MRI exposure and its disappearance immediately after MRI exposure. In the case of deflation failure, a second or third MRI exposure will be attempted, again followed by ultrasound confirmation of balloon deflation.

### Conventional reestablishment of the fetal airways

In the case of failure to deflate, balloon removal will be done as currently done with the conventional balloon, either ultrasound-guided puncture, fetoscopy, or in an emergency during abdominal delivery while the fetus is on placental circulation, or after birth by puncture above the manubrium sterni[15].

## ETHICS AND REGULATORY CONSIDERATIONS

In France, approval was provided by the committee for the protection of persons concerned (CPP “Ile de France VIII”) in January 2021 (# 21 01 01), and the French medicines controls authorities (ANSM) in March (2020-A02834-35-A). In Belgium, approval was given by the Ethics Committee on Clinical Studies of the University Hospitals Leuven in July 2021 (S65423). The study was registered at the Federal Agency for Medicines and Health Products (FAGG/80M0892).

## DISCUSSION

Based on robust clinical evidence, one should consider the option of FETO in selected fetuses with CDH [2-5]. One of the major concerns about FETO is the potential problems related to balloon removal [10]. The Smart-TO balloon addresses this issue by allowing a noninvasive, easily triggered, and externally controlled reversal of occlusion [16]. After extensive translational research, the time has come to assess the efficacy of reversal of the occlusion and the safety of this new device in a first-in-woman study.

The main objective of this study is to demonstrate the ability to successfully deflate the Smart-TO balloon by the magnetic fringe field generated by an MRI scanner. The present trial also aims to demonstrate the Smart-TO balloon is no longer within the airways. Non-visualization of the balloon will provide evidence for airway permeability. In the Belgian site, the E.C. also required to positively identify the localization of the balloon following deflation, either within the amniotic fluid, membranes, or placenta (at delivery), and exclude its persistence in the uterus by postpartum ultrasound. Additional objectives of this study include the evaluation of safety, even though no serious adverse effects directly related to the Smart-TO balloon are anticipated.

The dimensions of the Smart-TO balloon and material (latex) are the same as the Goldbal2® balloon. For this reason, it is anticipated that the Smart-TO will induce similar lung growth compared to the Goldbal2® balloon, as previously demonstrated in preclinical studies [12, 17]. Additional outcome measurements include the occurrence of membrane rupture and preterm delivery, which is consistently reported in all FETO series. We will also report on the consequence of the above.

The limitations of our study will be that this is a non-comparative trial. However, including a second arm, where controls would have FETO by means of the Goldbal2® balloon, appears to be unethical, since this will not provide new data and there is sufficient data on file on outcomes when using the standard balloon.

In conclusion, this first in-woman study aims to demonstrate the ability of Smart-TO balloon to be prenatally deflated by the magnetic fringe field generated by an MRI scanner, its expelling from the airways, as well as the safety of its use.

**Figure 1:**
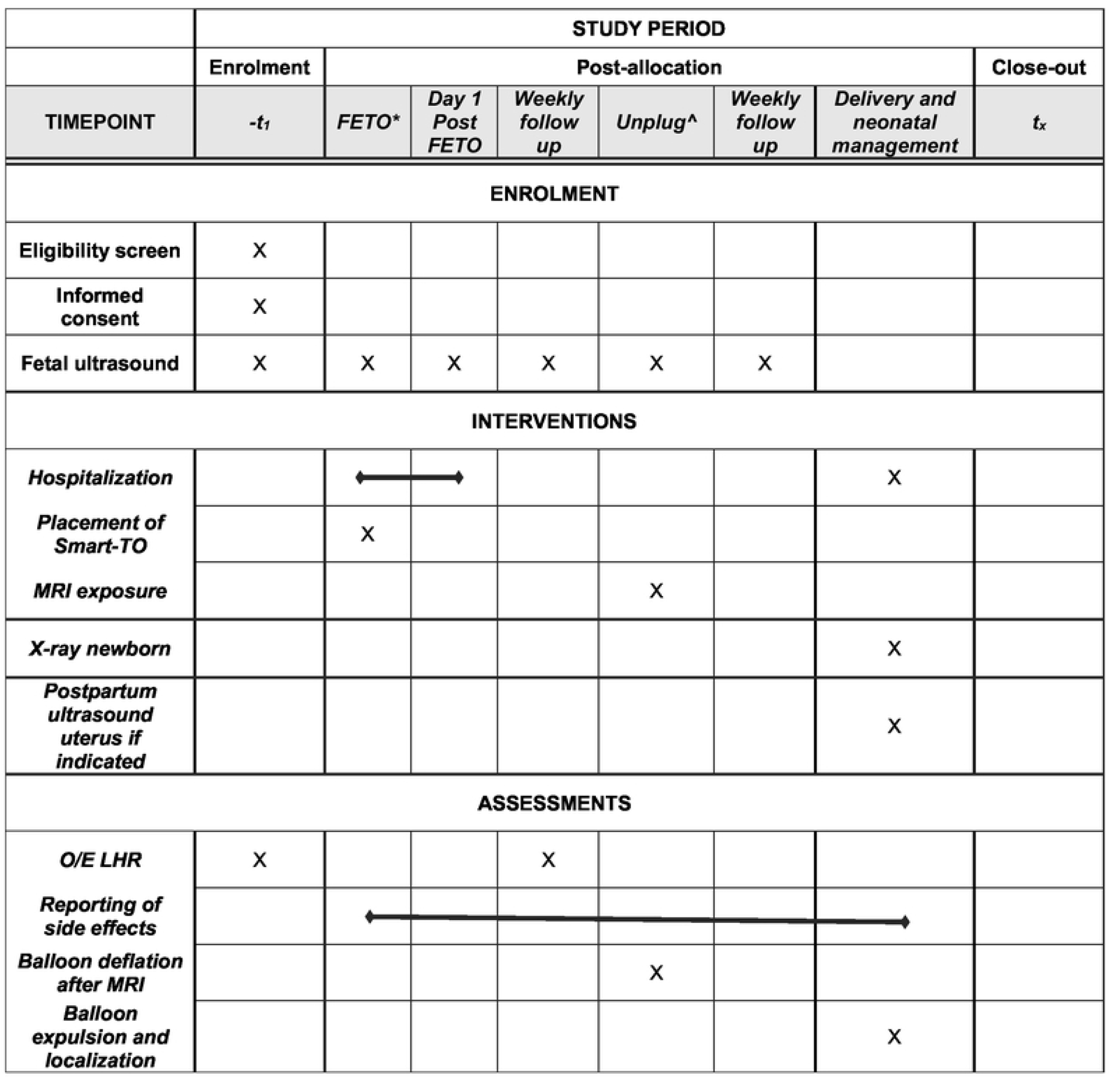
SPIRIT schedule. Abbreviations: FETO, Fetoscopic Endoluminal Tracheal Occlusion; O/E LHR, observed-to-expected lung-to-head ratio; MRI, Magnetic Resonance Image.

## Data Availability

No datasets were generated or analysed during the current study. All relevant data from this study will be made available upon study completion.

## Acknowledgements

The Smart-TO balloon is the result of several years of research and development within a consortium of the following institutions: BS-Medical Tech Industry (manufacturer), Strasbourg University Hospital, University of Strasbourg, INSERM Unit 1121 “Biomaterials and Bioengineering”, Institut Hospitalo-Universitaire de Strasbourg, Institute for Research Against Cancers of the Digestive System, SATT-Conectus Alsace, Simian Laboratory Europe, and the KU Leuven.

The authors thank URC-CIC Paris Centre (Adèle Bellino), the DRCI (Shoreh Azimi) and UZ Leuven Clinical Trial Centre (Veerle Doozen) for the implementation, monitoring and data management of the study.

## REFERENCES

1. Ameis, D., N. Khoshgoo, and R. Keijzer, Abnormal lung development in congenital diaphragmatic hernia. Semin Pediatr Surg, 2017. 26(3): p. 123–128.

2. Deprest, J.A., et al., Randomized Trial of Fetal Surgery for Moderate Left Diaphragmatic Hernia. N Engl J Med, 2021. 385(2): p. 119–129.

3. Deprest, J.A., et al., Randomized Trial of Fetal Surgery for Severe Left Diaphragmatic Hernia. N Engl J Med, 2021. 385(2): p. 107–118.

4. Russo, F.M., et al., Fetal endoscopic tracheal occlusion reverses the natural history of right-sided congenital diaphragmatic hernia: European multicenter experience. Ultrasound Obstet Gynecol, 2021. 57(3): p. 378–385.

5. Van Calster, B., et al., The randomized TOTAL-trials on fetal surgery for congenital diaphragmatic hernia: re-analysis using pooled data. Am J Obstet Gynecol, 2021.

6. Araujo Junior, E., et al., Erratum: Procedure-Related Complications and Survival Following Fetoscopic Endotracheal Occlusion (FETO) for Severe Congenital Diaphragmatic Hernia: Systematic Review and Meta-Analysis in the FETO Era. Eur J Pediatr Surg, 2017. 27(4): p. e1.

7. Araujo Junior, E., et al., Procedure-Related Complications and Survival Following Fetoscopic Endotracheal Occlusion (FETO) for Severe Congenital Diaphragmatic Hernia: Systematic Review and Meta-Analysis in the FETO Era. Eur J Pediatr Surg, 2017. 27(4): p. 297–305.

8. Deprest, J., et al., Technical aspects of fetal endoscopic tracheal occlusion for congenital diaphragmatic hernia. Journal of pediatric surgery, 2011. 46(1): p. 22–32.

9. Done, E., et al., Predictors of neonatal morbidity in fetuses with severe isolated congenital diaphragmatic hernia undergoing fetoscopic tracheal occlusion. Ultrasound Obstet Gynecol, 2013. 42(1): p. 77–83.

10. Jimenez, J.A., et al., Balloon removal after fetoscopic endoluminal tracheal occlusion for congenital diaphragmatic hernia. Am J Obstet Gynecol, 2017.

11. Sananes, N., et al., Evaluation of a new balloon for fetal endoscopic tracheal occlusion in the nonhuman primate model. Prenat Diagn, 2019.

12. Basurto, D., et al., New device permitting non-invasive reversal of fetal endoscopic tracheal occlusion: ex-vivo and in-vivo study. Ultrasound Obstet Gynecol, 2020. 56(4): p. 522–531.

13. Basurto, D., et al., Safety and efficacy of the Smart Tracheal Occlusion device in the diaphragmatic hernia lamb model. Ultrasound Obstet Gynecol, 2020.

14. Vollset, S.E., Confidence intervals for a binomial proportion. Stat Med, 1993. 12(9): p. 809–24.

15. Van der Veeken, L., et al., Fetoscopic endoluminal tracheal occlusion and reestablishment of fetal airways for congenital diaphragmatic hernia. Gynecol Surg, 2018. 15(1): p. 9.

16. Sananes, N., et al., Evaluation of a new balloon for fetal endoscopic tracheal occlusion in the non-human primate model. Prenat Diagn, 2019.

17. Basurto, D., et al., Safety and efficacy of smart tracheal occlusion device in diaphragmatic hernia lamb model. Ultrasound Obstet Gynecol, 2021. 57(1): p. 105–112.

